# Estimation of Oral Disease Burden from Claims and Self-Reported Data

**DOI:** 10.1101/2021.07.01.21259799

**Authors:** Christopher Okunseri, Julie Frantsve-Hawley, Madhuli Thakkar-Samtani, Ilya Okunev, Eric P. Tranby

## Abstract

**Objective:** To examine the use of Medicaid, commercial claims, and self-reported survey data to estimate the prevalence of oral disease burden.

**Methods:** We analyzed 2018 Medicaid claims from IBM Watson Medicaid Marketscan database, commercial claims from the IBM Dental Database, and Medical Expenditure Panel Survey (MEPS) data. The estimate of oral disease burden was based on standard metrics using periodontal and caries-related treatment procedure codes. Examples are restorations: D2000-D2999, root Canals: D3230 – D3334, periodontics: D4000 – D4999, prosthodontics: D5000 – D6999 and extractions: D7000 - D7251. A direct comparison between the data sets was also done. Enrollees from the different databases were broken down by gender, race/ethnicity, and into age groups.

**Results:** Medicaid and commercial enrollees were 11.6 million and 10.5 million. The weighted proportion from MEPS for Medicaid and commercial plans ranged from 80-208 million people. Prevalence of caries-related treatments was estimated for IBM Watson and MEPS for total enrollees for Medicaid (13% vs. 12%); and commercial claims (25% vs. 17%), respectively. Prevalence of periodontal related treatments was estimated for IBM Watson and MEPS total enrollees for Medicaid (0.7% vs. 0.5%) and commercial claims (7% vs. 1.6%), respectively. Prevalence of dental diseases was higher in patients with at least one visit for Medicaid, commercial plans, and MEPS. Prevalence based on specific procedures were higher in commercial plans than in Medicaid.

**Conclusions:** Claims data has the potential to serve as a proxy measure for the estimate of dental disease burden in a population. In addition, in rare events, claims data provides a better estimate of disease burden because it is based on a larger dataset.

## INTRODUCTION

To estimate prevalence for a condition, researchers would seek out the opportunity to conduct an epidemiological survey with the goal to randomly select a reasonable sample size from the entire population. This entails counting the number of persons affected by the disease in a geographic location at a specified time to provides prevalence estimates for the condition. However, challenges exist in conducting epidemiological surveys such as cost, time, and logistics. In comparison, administrative claims data are readily available for different subgroups, cover a large proportion of a specified population, and is collected in the same manner over time. Therefore, claims data could serve as an alternative source to provide valid measure of disease burden (1).

Administrative claims from commercial and public health insurance plans provide a rich source of information for planning and improving healthcare delivery systems. Health service researchers use claims data to capture charges/costs, quality, access, and utilization of health services. Administrators use claims for billing, payments, fraud identification, and to define health benefits as well as to identify enrollees who are at higher risk of needing more services (1). Administrative claims data have been used to estimate prevalence and incidence of systemic diseases (2-8). For example, claims data were used to determine prevalence of hypertension, but with some inherent variability in sensitivity and specificity values (8). Additionally, Craign et al determine prevalence of a cluster of sarcoidosis in Vermont and authors observed that the prevalence of the disease exceeded limited and unsubstantiated U.S. reports (4). Sloan et al estimated the incidence rate of chronic eye disorders from longitudinal Medicare claims data. Findings from the study were comparable to those observed from population-based studies. (7).

Despite the availability of these studies, the use of dental claims to estimate prevalence of dental disease burden is understudied, is poorly understood, and is almost non-existent or limited in scope. This in part can be attributed to the complexity of the analytical approach and interpretation of findings with its associated limitations. Such as the lack of diagnostic codes not been widely used and promoted in dentistry. Traditionally, dental Medicaid and commercial claims provide information on the financial expenditures associated with dental treatment (9-11) and self-reported measures documents individual’s behavior, attitude, symptoms, and treatment received (12-13). However, commercial and Medicaid claims data built on Current Dental Terminology (CDT) procedure codes could potentially be used to provide estimates on the prevalence for oral disease such as dental caries and periodontal disease in sub-populations who have dental coverage.

In this study, we examined the use of Medicaid and commercial claims data to estimate the burden of oral disease and compare disease prevalence rates in claims and survey data. We use CDT procedure codes to estimate the prevalence of dental caries and periodontal disease in a sub-population of Medicaid and commercial beneficiaries with dental coverage. To ascertain the validity and reliability of estimates, we conducted comparative analysis on claims data and self-reported survey data on dental service utilization for similar sub-populations. To our knowledge, no study has analyzed data from these discrete datasets to estimate the burden of dental disease.

## METHODS

Medicaid and commercial claims data from IBM Watson Marketscan Databases for 2018 as well as publicly available and nationally representative survey data from the 2018 Medical Expenditure Panel Study (MEPS) were analyzed for this study. Patients’ claim history in the 2018 IBM Watson Medicaid Multi-State Marketscan Database and the IBM Watson Dental Database were used to estimate the prevalence of oral disease (See Appendix 1 for full inclusion criteria and variable definitions). The Medicaid dataset contains enrollment, medical and dental claims information from 13 de-identified states. The dental database contains claims from commercially insured patients and includes enrollment, medical and dental claims information from select insurance plans in all 50 states. From this data, we divided enrollees into 3 age groups: 0-20, 21-64, 65-90+. These categories reflect in part Medicaid coverage of routine dental care, which is mandatory for children 20 and under, but optional for adults 21 and over, with significant variation in coverage across states. We also stratified patients by gender (male or female) as well as by race/ethnicity available in the Medicaid data (White, Black, Hispanic, and Other). However, race/ethnicity information were not available in the commercially insured population. We focused on 5 procedure code groupings in our analysis for the burden of oral disease: Restorative care = D2000 – D2999, Endodontics = D3230 – D3334, Periodontics = D4000 – D4999, Prosthodontics = D5000 – D6999 and Extractions = D7000 – D7251. Patients were deemed to have caries if they received a restorative care, endodontic procedure, prosthodontics, or oral surgery (CDT codes D2000 – D3999, D5000 – D7999), and were assumed to have periodontal disease if they had a code associated with the disease.

Self-reported treatment for oral disease was extracted from the Medical Expenditure Panel Survey (MEPS) for Medicaid enrollees and for those with commercial insurance for 2018. The MEPS is a nationally representative survey conducted by the U.S. Agency for Healthcare Research and Quality (AHRQ) and the National Center for Health Statistics (NCHS). This ongoing survey is designed to collect information on healthcare utilization, medical expenditures, and insurance coverage as well as various socioeconomic and demographic characteristics for the U.S. civilian and non-institutionalized population. We identified Medicaid enrollees as respondents who answered yes to ever having Medicaid/SCHIP coverage in 2018. Similarly, commercially insured respondents were identified as individuals who reported yes to ever having private insurance in 2018. We examined five self-reported measures for oral disease—restorations defined as any fillings, inlays, crowns, endodontic procedures defined as having any root canals; extractions defined as having any tooth pulled or other oral surgery; prosthodontic procedures defined as having any fixed prosthesis or relining/repair of bridges/dentures or implants; and periodontics defined as having any periodontal scaling, root planning or gum surgery. This information was stratified by Medicaid enrollees and those commercially insured. We further estimated prevalence of oral disease in a sub-population of individuals with at least one dental visit in 2018.

Based on the data, we produced estimates of the number and proportion of individuals with reported caries-related procedures and periodontal disease related procedures by age, gender and race/ethnicity among those enrolled in the different insurance programs. In this study, we also compared estimates from these different sources to understand how estimates of the burden for disease could depend on the data collection method, the quality and completeness of the data, and the representativeness of the data.

## RESULTS

Table 1 shows the study population from 3 datasets in 2018. Medicaid claims from IBM Watson Marketscan Multi-State Marketscan Database, commercial claims from IBM Watson Dental Database and weighted survey data from Medical Expenditure Panel Survey. Overall, the number of Medicaid enrollees in the IBM Watson data was 11.5 million and, in the MEPS data was 79.5 million. The commercially insured enrollees were 10.5 million in the IBM Watson data and 208 million in the MEPS data. The MEPS survey are weighted estimates of the population derived from a sample of 8,767 covered by Medicaid and 17,565 covered by commercial insurance (See Appendix 2). On the other hand, the claims data represents the entire universe of enrollees.

**Table 1:**
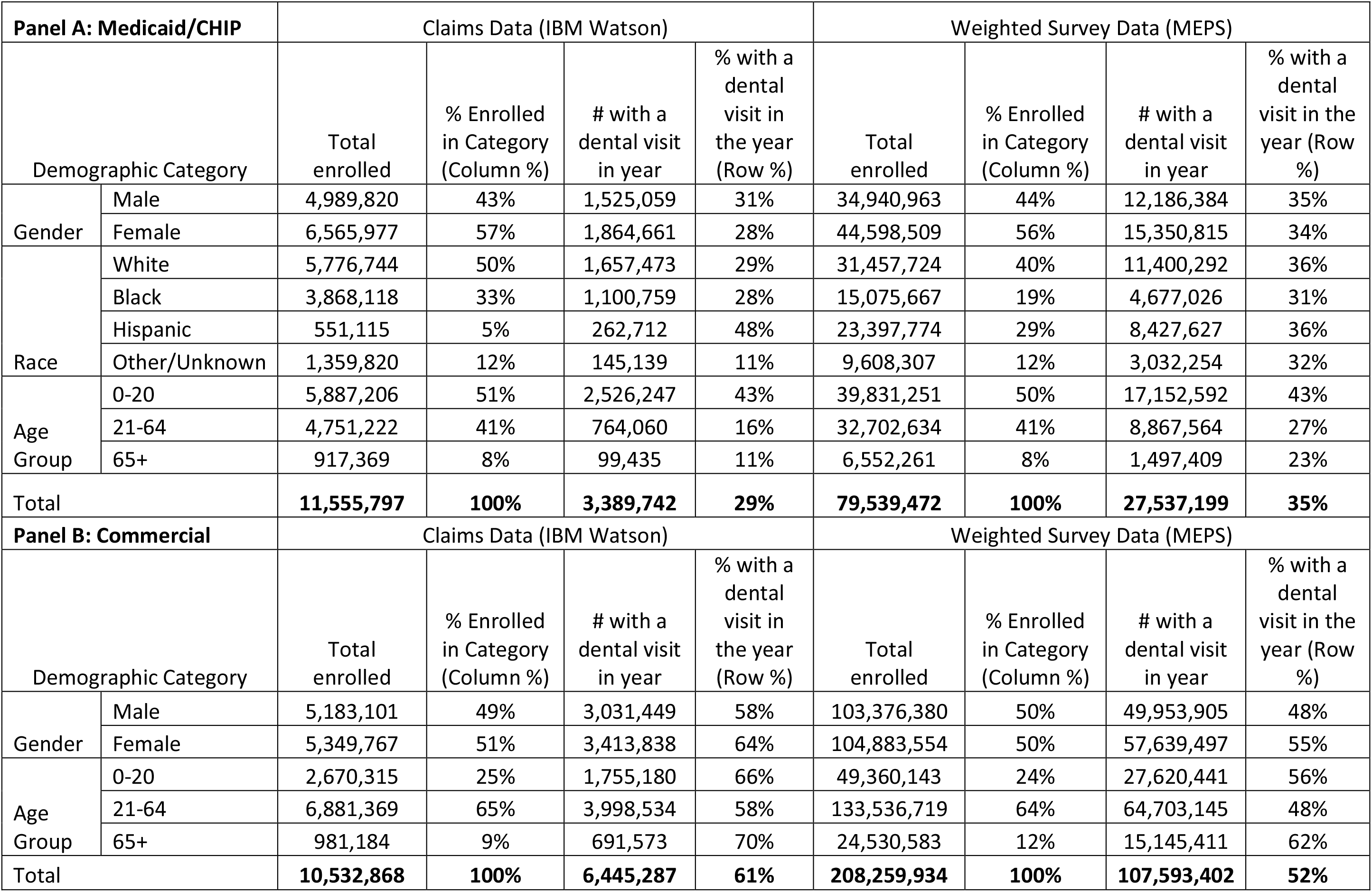
Study Populations Across Two Datasets for The Estimation of Dental Disease Burden.

Overall, the number of Medicaid enrollees with a dental visit was 3.4 million for the IBM Watson Medicaid dataset and 27.5 million for MEPS Medicaid population. Among those covered by commercial dental plans, 6.5 million and 1.8 million had a dental visit in 2018, respectively. The data sources provided similar windows in the demographic distribution of enrollees. Approximately, 51% vs. 50% were aged 0-20 in the Medicaid claims and weighted MEPS data respectively. Similarly, 25% vs. 24% of enrollees were aged 0-20 in the commercial claims and weighted MEPS data, respectively. This age distribution is reflective of the Medicaid population in the United States (14). Among Medicaid enrollees, a higher percentage of them were females (57% in the claims data and 56% in the survey data). The datasets vary in their enrollee distributions by race/ethnicity. Half of the enrolled population in the Medicaid claims data were White compared to 40% in the survey data. The Black population ranged from a high of 33% in the Medicaid claims data to a low of 19% in the survey data. Meanwhile, the Hispanic population was largest in the survey data (29%) and smallest in the claims data (5%). Among those insured by commercial dental plans, there were similar proportions of both males and females. About a quarter of the participants in the commercial datasets were 0-20 years, slightly less than two thirds were working age adults, 21 to 64 (65% in claims, and 64% in the survey) and 65+ (9%, and 12%) respectively.

Estimates of the proportion of the population with a dental visit in 2018 vary widely by data source. Among those enrolled in Medicaid, 35% of the enrolled population self-reported a dental visit in the 2018 survey data, but 29% visited a dentist in the claims data. Among those in commercial dental plans, 52% reported a dental visit in 2018 survey data, while 61% had a claim related to a dental visit. Among those enrolled in Medicaid, males had slightly higher rates of dental visits than the overall average, while females had slightly lower rates. The situation is reversed among those commercially insured. Reflective of policy coverage differences, children were the most likely to have a dental visit in 2018 in Medicaid data sources at 43%. Medicaid enrolled adults were much less likely to have a dental visit in 2018 than children, with estimates ranging from 16% in the claims data and 27% in the survey data, and those 65+ were the least likely to have a dental visit. In the commercially insured population, those 65+ had the highest rates of dental visits, followed by children 20 and under, and then working age adults. Substantial racial differences were observed across Medicaid datasets, with Hispanics having the highest rates of dental visits in the IBM claims data at 48% and being closer to the population average in the other datasets. On the other hand, Blacks had dental visit rates below the population average in the datasets.

Table 2, Panel A shows the dental procedures received for treatment for caries and periodontal disease from the IBM claims and self-reported MEPS survey databases among Medicaid enrollees. Gender was not shown in these tables because there was very little variation by gender. The prevalence of caries related treatment was almost identical across both datasets for total enrollees, with IBM Watson at 13% and MEPS at 12%. However, caries was more prevalent among dental patients in the claims data (43%) than in the survey data (36%). In Table 2, Panel B, showing information on commercially insured patients, caries was more prevalent among both enrollees and dental patients in the claims data (25% and 40%, respectively) than in the MEPS survey data (17% among enrollees and 33% among dental patients).

**Table 2:** Treatment for Caries and Periodontal Disease in Claims Data and Self-Reported Survey Data.

| Panel A: Medicaid/CHIP |  |  |  |  |  |  |  |  |  |  |  |  |  |
| --- | --- | --- | --- | --- | --- | --- | --- | --- | --- | --- | --- | --- | --- |
|  |  | Caries Treatment<br>(Restorations, Root Canals, Extractions, Prosthodontics) |  |  |  |  |  | Periodontal Treatment |  |  |  |  |  |
|  |  | Count |  | Prevalence among<br>Total Enrolled |  | Prevalence among<br>Dental Patients |  | Count |  | Prevalence<br>among Total<br>Enrolled |  | Prevalence<br>among Dental<br>Patients |  |
|  |  | Claims<br>(IBM<br>Watson) | Survey<br>(MEPS) | Claims<br>(IBM<br>Watson) | Survey<br>(MEPS) | Claims<br>(IBM<br>Watson) | Survey<br>(MEPS) | Claims<br>(IBM<br>Watson) | Survey<br>(MEPS) | Claims<br>(IBM<br>Watson) | Survey<br>(MEPS) | Claims<br>(IBM<br>Watson) | Survey<br>(MEPS) |
| Race | White | 744,790 | 4,199,409 | 13% | 13% | 45% | 37% | 38,046 | 191,832 | 0.7% | 0.6% | 2.3% | 1.7% |
|  | Black | 444,023 | 1,761,880 | 11% | 12% | 40% | 38% | 32,388 | 83,703 | 0.8% | 0.6% | 2.9% | 1.8% |
|  | Hispanic | 103,044 | 2,874,408 | 19% | 12% | 39% | 34% | 4,698 | 51,837 | 0.9% | 0.2% | 1.8% | 0.6% |
|  | Other | 60,044 | 1,072,764 | 4% | 11% | 41% | 35% | 3,756 | 49,700 | 0.3% | 0.5% | 2.6% | 1.6% |
| Age<br>Group | 0-20 | 924,184 | 4,184,853 | 16% | 11% | 37% | 24% | 11,741 | 37,209 | 0.2% | 0.1% | 0.5% | 0.2% |
|  | 21-64 | 472,783 | 4,753,922 | 10% | 15% | 62% | 54% | 65,435 | 332,635 | 1.4% | 1.0% | 8.6% | 3.8% |
|  | 65+ | 47,637 | 958,339 | 5% | 15% | 48% | 64% | 6,103 | 7,227 | 0.7% | 0.1% | 6.1% | 0.5% |
| Total |  | **1,444,604** | **9,908,462** | **13%** | **12%** | **43%** | **36%** | **83,279** | **377,072** | **0.7%** | **0.5%** | **2.5%** | **1.4%** |
| Panel B: Commercial |  |  |  |  |  |  |  |  |  |  |  |  |  |
|  |  | Caries Treatment<br>(Restorations, Root Canals, Extractions, Prosthodontics) |  |  |  |  |  | Periodontal Treatment |  |  |  |  |  |
|  |  | Count |  | Prevalence among<br>Total Enrolled |  | Prevalence among<br>Dental Patients |  | Count |  | Prevalence<br>among Total<br>Enrolled |  | Prevalence<br>among Dental<br>Patients |  |
|  |  | Claims | Survey | Claims | Survey | Claims | Survey | Claims | Survey | Claims | Survey | Claims | Survey |
| Age<br>Group | 0-20 | 508,487 | 5,321,277 | 19% | 11% | 29% | 19% | 11,131 | 173,243 | 0.4% | 0.4% | 0% | 1% |
|  | 21-64 | 1,734,451 | 22,664,178 | 25% | 17% | 43% | 35% | 578,789 | 2,494,241 | 8% | 2% | 14% | 4% |
|  | 65+ | 356,396 | 7,191,316 | 36% | 29% | 52% | 47% | 126,974 | 647,326 | 13% | 3% | 18% | 4% |
| Total |  | **2,599,334** | **35,188,119** | **25%** | **17%** | **40%** | **33%** | **716,894** | **3,314,811** | **7%** | **1.6%** | **11%** | **3%** |

In both Medicaid and commercially insured populations, prevalence of periodontal disease was relatively low and substantially below the estimates of the burden of disease from surveillance data (11) on periodontitis (Table 2). In Medicaid, the prevalence of periodontal disease was slightly higher in the IBM Watson claims data at 0.7%, then in the MEPS at 0.5%. Among commercially insured patients, prevalence was 7% in the claims data and only 1.6% based on self-reports. Similarly, periodontal disease was more prevalent among dental patients in the claims data (2.5% in Medicaid and 11% in Commercial) than in the survey data (1.4% in Medicaid and 3% in commercial). In every age group, we observed a higher prevalence of periodontal disease from claims data than in the survey estimates. Periodontal disease prevalence was highest among adults, with very small prevalence among children.

Table 3a shows caries-related dental procedures among Medicaid total enrolled and dental enrollees and by self-reported data. The prevalence of restorative care and extractions was higher in the claims data than in in the survey data. On the other hand, the prevalence of root canals and prosthodontics was higher in the survey data than in the claims data.

**Table 3a:**
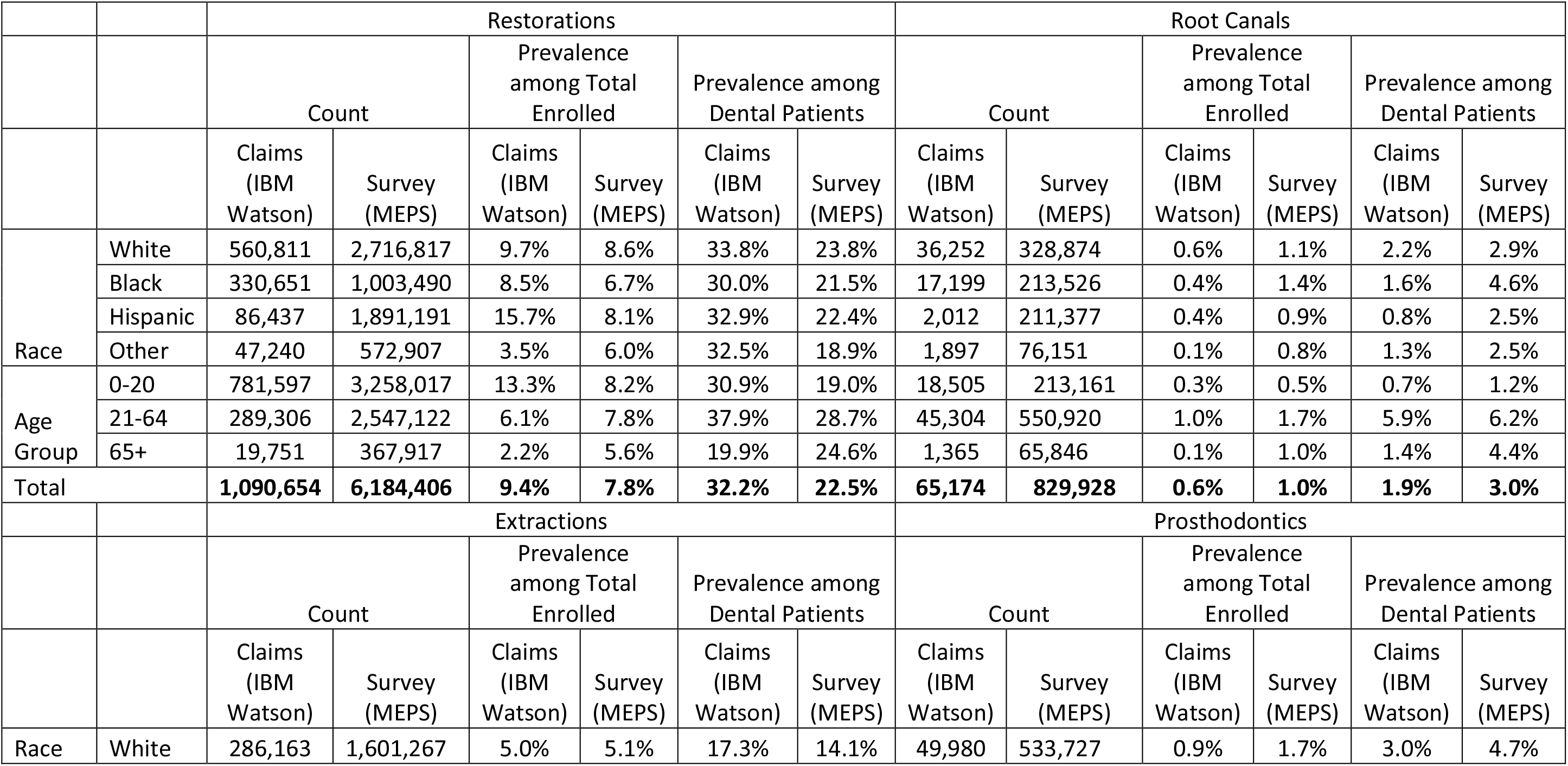
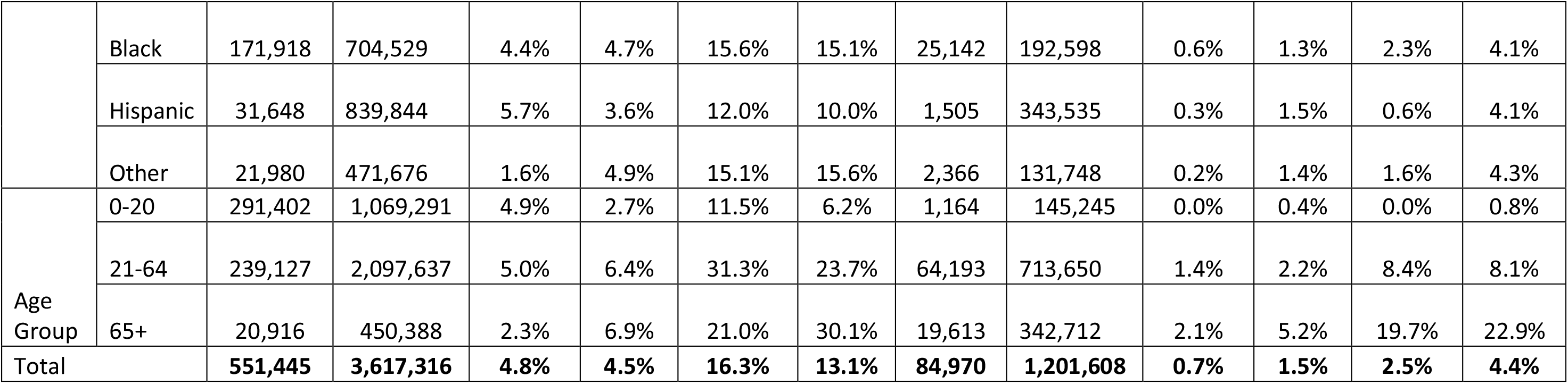
Medicaid Enrolled Population Receiving Dental Care in Categories in Claims Data and Self-Reported Survey Data.

|  |  | Restorations |  |  |  |  |  | Root Canals |  |  |  |  |  |
| --- | --- | --- | --- | --- | --- | --- | --- | --- | --- | --- | --- | --- | --- |
|  |  | Count |  | Prevalence among Total Enrolled |  | Prevalence among Dental Patients |  | Count |  | Prevalence among Total Enrolled |  | Prevalence among Dental Patients |  |
|  |  | Claims (IBM Watson) | Survey (MEPS) | Claims (IBM Watson) | Survey (MEPS) | Claims (IBM Watson) | Survey (MEPS) | Claims (IBM Watson) | Survey (MEPS) | Claims (IBM Watson) | Survey (MEPS) | Claims (IBM Watson) | Survey (MEPS) |
| Race | White | 560,811 | 2,716,817 | 9.7% | 8.6% | 33.8% | 23.8% | 36,252 | 328,874 | 0.6% | 1.1% | 2.2% | 2.9% |
|  | Black | 330,651 | 1,003,490 | 8.5% | 6.7% | 30.0% | 21.5% | 17,199 | 213,526 | 0.4% | 1.4% | 1.6% | 4.6% |
|  | Hispanic | 86,437 | 1,891,191 | 15.7% | 8.1% | 32.9% | 22.4% | 2,012 | 211,377 | 0.4% | 0.9% | 0.8% | 2.5% |
|  | Other | 47,240 | 572,907 | 3.5% | 6.0% | 32.5% | 18.9% | 1,897 | 76,151 | 0.1% | 0.8% | 1.3% | 2.5% |
| Age Group | 0-20 | 781,597 | 3,258,017 | 13.3% | 8.2% | 30.9% | 19.0% | 18,505 | 213,161 | 0.3% | 0.5% | 0.7% | 1.2% |
|  | 21-64 | 289,306 | 2,547,122 | 6.1% | 7.8% | 37.9% | 28.7% | 45,304 | 550,920 | 1.0% | 1.7% | 5.9% | 6.2% |
|  | 65+ | 19,751 | 367,917 | 2.2% | 5.6% | 19.9% | 24.6% | 1,365 | 65,846 | 0.1% | 1.0% | 1.4% | 4.4% |
| Total |  | **1,090,654** | **6,184,406** | **9.4%** | **7.8%** | **32.2%** | **22.5%** | **65,174** | **829,928** | **0.6%** | **1.0%** | **1.9%** | **3.0%** |
|  |  | Extractions |  |  |  |  |  | Prosthodontics |  |  |  |  |  |
|  |  | Count |  | Prevalence among Total Enrolled |  | Prevalence among Dental Patients |  | Count |  | Prevalence among Total Enrolled |  | Prevalence among Dental Patients |  |
|  |  | Claims (IBM Watson) | Survey (MEPS) | Claims (IBM Watson) | Survey (MEPS) | Claims (IBM Watson) | Survey (MEPS) | Claims (IBM Watson) | Survey (MEPS) | Claims (IBM Watson) | Survey (MEPS) | Claims (IBM Watson) | Survey (MEPS) |
| Race | White | 286,163 | 1,601,267 | 5.0% | 5.1% | 17.3% | 14.1% | 49,980 | 533,727 | 0.9% | 1.7% | 3.0% | 4.7% |
|  | Black | 171,918 | 704,529 | 4.4% | 4.7% | 15.6% | 15.1% | 25,142 | 192,598 | 0.6% | 1.3% | 2.3% | 4.1% |
|  | Hispanic | 31,648 | 839,844 | 5.7% | 3.6% | 12.0% | 10.0% | 1,505 | 343,535 | 0.3% | 1.5% | 0.6% | 4.1% |
|  | Other | 21,980 | 471,676 | 1.6% | 4.9% | 15.1% | 15.6% | 2,366 | 131,748 | 0.2% | 1.4% | 1.6% | 4.3% |
| Age Group | 0-20 | 291,402 | 1,069,291 | 4.9% | 2.7% | 11.5% | 6.2% | 1,164 | 145,245 | 0.0% | 0.4% | 0.0% | 0.8% |
|  | 21-64 | 239,127 | 2,097,637 | 5.0% | 6.4% | 31.3% | 23.7% | 64,193 | 713,650 | 1.4% | 2.2% | 8.4% | 8.1% |
|  | 65+ | 20,916 | 450,388 | 2.3% | 6.9% | 21.0% | 30.1% | 19,613 | 342,712 | 2.1% | 5.2% | 19.7% | 22.9% |
| Total |  | **551,445** | **3,617,316** | **4.8%** | **4.5%** | **16.3%** | **13.1%** | **84,970** | **1,201,608** | **0.7%** | **1.5%** | **2.5%** | **4.4%** |

Table 3b shows the caries-related dental procedures among enrollees in commercial dental plans. The prevalence of restorative care and extractions were higher in the claims data than in the survey data, and the prevalence of root canals and prosthodontics having similar estimates between the two types of data or higher in the survey data. those enrolled in Medicaid.

**Table 3b:** Commercially Insured Population Receiving Dental Care in Categories in Claims Data and Self-Reported Survey Data.

|  |  | Restorations |  |  |  |  |  | Root Canals |  |  |  |  |  |
| --- | --- | --- | --- | --- | --- | --- | --- | --- | --- | --- | --- | --- | --- |
|  |  | Count |  | Prevalence among Total Enrolled |  | Prevalence among Dental Patients |  | Count |  | Prevalence among Total Enrolled |  | Prevalence among Dental Patients |  |
|  |  | Claims | Survey | Claims | Survey | Claims | Survey | Claims | Survey | Claims | Survey | Claims | Survey |
| Age Group | 0-20 | 387,648 | 3,585,212 | 14.5% | 7.3% | 22.1% | 13.0% | 13,251 | 177,225 | 0.5% | 0.4% | 0.8% | 0.6% |
|  | 21-64 | 1,462,290 | 17,138,019 | 21.2% | 12.8% | 36.6% | 26.5% | 198,885 | 2,834,886 | 2.9% | 2.1% | 5.0% | 4.4% |
|  | 65+ | 286,808 | 4,947,158 | 29.2% | 20.2% | 41.5% | 32.7% | 34,414 | 658,219 | 3.5% | 2.7% | 5.0% | 4.3% |
| Total |  | **2,136,746** | **25,681,738** | **20.3%** | **12.3%** | **33.2%** | **23.9%** | **246,550** | **3,670,330** | **2.3%** | **1.8%** | **3.8%** | **3.4%** |
|  |  | Extractions |  |  |  |  |  | Prosthodontics |  |  |  |  |  |
|  |  | Count |  | Prevalence among Total Enrolled |  | Prevalence among Dental Patients |  | Count |  | Prevalence among Total Enrolled |  | Prevalence among Dental Patients |  |
|  |  | Claims | Survey | Claims | Survey | Claims | Survey | Claims | Survey | Claims | Survey | Claims | Survey |
| Age Group | 0-20 | 162,702 | 1,750,378 | 6.1% | 3.5% | 9.3% | 6.3% | 2,166 | 252,169 | 0.1% | 0.5% | 0.1% | 0.9% |
|  | 21-64 | 308,531 | 4,510,956 | 4.5% | 3.4% | 7.7% | 7.0% | 158,535 | 2,388,553 | 2.3% | 1.8% | 4.0% | 3.7% |
|  | 65+ | 70,803 | 1,662,597 | 7.2% | 6.8% | 10.2% | 11.0% | 71,403 | 1,787,634 | 7.3% | 7.3% | 10.3% | 11.8% |
| Total |  | **542,036** | **7,923,933** | **5.1%** | **3.8%** | **8.4%** | **7.4%** | **232,104** | **4,428,356** | **2.2%** | **2.1%** | **3.6%** | **4.1%** |

## DISCUSSION

### Summary of main results and their interpretation

This study used three different datasets to estimate the burden of dental disease. Although claims data have not been previously utilized to estimate the prevalence of dental disease, our comparative study suggests that claims data can be just as valuable as survey data when it comes to capturing burden of dental disease. We observed that the dental access rates varied by claims and survey data sources. The observed difference in dental visits likely reflect differences in sampling type and the proxy measure used to capture dental disease burden. On the other hand, estimates from claims data have a high degree of internal reliability. Rare events, such as the prevalence of periodontal treatment among enrollees, are better estimated in claims data, due to the large size of the datasets. Estimates of these events in survey data are more prone to random variation with 1 or 2 cases substantially changing estimates of the burden of the disease in weighted data, as can be seen among children (Appendix 2).

Among Medicaid and commercial enrollees, the burden of dental disease estimates across multiple measures were higher in claims than in survey data. The observed difference in the burden of dental disease from claims data could have resulted from the type of insurance coverage, use of dental services and access to care of the enrollees. Information from claims data is somewhat generalizable because it is from a larger universe of Medicaid and commercial enrollees even without matching them to diagnostic codes. Claims data have the potential to project future dental needs within sub-populations that have insurance coverage and to capture oral disease status were prevalence rates from survey data is low, such as periodontal disease in children or oral cancers in adults.

For caries-related dental procedures for Medicaid and commercial enrollees, we observed a substantial difference in the prevalence estimates in the burden of dental disease depending on whether we used total enrollees or dental patients (dental enrollees with at least one documented dental visit) for the denominator. Total enrollees could potentially represent those with treated and untreated caries even though we do not know the level of untreated caries among enrollees that did not have access to care. Dental patients represent mostly those with treated disease as seen from the claims data. Although, our study did not attempt to validate this assumption, the difference in proportion helps to somewhat justify our assumption.

In terms of age, total enrolled population estimates for the burden of dental disease identified by caries-related procedures progressively decreased with age in Medicaid claims, but increased with age in commercial claims. The trend seen with Medicaid claims could have resulted from the lack of adult dental benefits experienced in some states. However, an increase with age was observed in the survey data for both Medicaid and commercial claims. This finding is consistent with what is typically reported from epidemiological surveys. For example, prevalence of total dental caries among youths increased from 50.5% to 53.8% among 6-11 years old and 12-19 years old (15).

In terms of specific caries-related procedures, claims data showed higher prevalence rates of restorative care and extraction services, though root canals and prosthodontics were more prevalent in survey data among Medicaid enrollees. However, these are relatively rare events in the survey data and like periodontal disease, these estimates may be prone to random variations. Among those insured with Medicaid and commercial dental plans, the prevalence of each procedure category by age are consistent with clinical expectations, the course of caries and the policy coverages. Among all age groups, restorations are the most common treatments for caries, followed by extractions and root canals, with prosthodontics being relatively uncommon.

Restorations were most prevalent among children in Medicaid while being most prevalent among seniors in commercial plans. Root canals and extractions were most common among working age adults in Medicaid and most prevalent among seniors in commercial. The estimates seen in the total enrolled population is assumed to represent the lower limit of disease burden and that for dental patients as the upper limit of dental disease burden. Prosthodontics were concentrated among seniors in all data sources. The prevalence of restorations was similar in both Medicaid and commercially insured populations. Extractions were slightly more common in Medicaid enrolled patients. Even though the primary purpose for the various datasets were somewhat different, the proportion of participants by gender and age group were almost the same in this descriptive study. Survey data are designed to provide population-based estimates. In addition, Medicaid and commercial claims data are representative of the enrollees in the program.

### Importance of results

The advantages of using claims data for this purpose include-1) it contains the entire universe of data among enrollees; 2) it is available in real-time; and 3) they are collected as part of standard business processes and as such does not require additional costs and effort to obtain the data, as is the case with survey and exam data. In addition, for rare events claims data provides a better estimate of disease because it is based on larger dataset. Survey data remain useful in estimating burden of disease especially when information from those not accessing care is critical.

The estimates from claims data also match well with other existing utilization and prevalence measures. Data from the annual Early and Periodic Screening, Diagnostic, and Treatment (EPSDT) Participation Report in 2018 revealed that 47% of children aged 0-20 years and enrolled in Medicaid received any dental care and 20% received dental treatment services. This can be correlated to our study’s estimates of the proportion from the claims data, with 43% of children enrolled in Medicaid having received a dental service and 16% receiving treatment for caries or periodontal care (16).

### Shortcomings and strengths of the study

Certain strengths and limitations must be considered when interpreting our findings. Claims data are internally and highly reliable because they contain the entire universe of observed information on the enrollee. However, they may underestimate the burden of disease because they rely on the dentist submitting a claim. Medicaid claims data are heavily influenced by policy and coverage limitations, which may impact their reliability in assessing certain disease states. This is especially true for adults for whom dental coverage is not mandated by law. Commercial claims data is somewhat restricted to those considered to be healthier, higher income and educational status. However, it covers close to 50% of the insurance dental population (17). Due to the differences in coverage between Medicaid and commercial insurance, one would expect that claims related to certain diseases will be more prevalent.

Self-reported survey data has less reliability as they rely on unbiased sampling and on individual’s reporting accurately and completely for accurate estimation. However, they allow the capture of information from patients who are seen in non-traditional settings or who do not have routine access to care. Both data are snapshots in time and allow the estimation of disease. In addition, the differences observed are likely due to the comprehensiveness of claims data, which incorporates everything done to the patient each year, whereas self-reported surveys may have a more limited view due to the limited number of questions, recall/recency bias, or a lack of knowledge of dental terminology by respondents.

### Future directions

We expect future studies to address some of the limitations identified in this study and propose new paradigm for measuring prevalence of oral disease burden from claims data. In addition, the general availability and details in claims data should be combined with International Classification of Diseases, 9^th^ or 10^th^ Revision, Clinical Modification (ICD-9-CM) diagnosis and procedure codes to provide relatively accurate information for understanding disease prevalence in a population (17). While some dentists are utilizing diagnostic codes, the usage is not required by most payers or agencies. Moreover, dentists are not routinely trained to use diagnostic codes, so continuing education and calibration will be required before widescale adoption.

### Public Health implications

Claim data have the potential to serve as proxy for the estimation of dental disease burden in a population. In addition, in rare events, claims data provides a better estimate of disease burden because it is based on a larger dataset. However, survey data is particularly useful when information from those not accessing care, which is not captured in claims data, is critical.

## Conclusion

The comparison of Medicaid and commercial claims data to self-reported survey data confirms that dental claims data can serve as proxy for estimating burden of disease in a population.

## Data Availability

The IBM Watson data is available a data use agreement. The nationally representative survey data from the 2018 Medical Expenditure Panel Study (MEPS) is publicly available.

## Appendix 1: Inclusion Criteria and Variable Definitions

### IBM Watson (Claims)

- Enrolled Population
  ∘ Included all patients enrolled for at least one day in the year 2018.
- Dental Patients
  ∘ Dental patients are defined as having at least one dental service from a dental provider in 2018. A dental service is defined by the presence of a CDT proc Code between D0001 and D9999.
- Procedure Groupings as defined by CDT procedure codes
  ∘ Restorations: D2000-D2999
  ∘ Root Canals: D3230 – D3334
  ∘ Periodontics: D4000 – D4999
  ∘ Prosthodontics: D5000 – D6999
  ∘ Extractions: D7000 - D7251
- Definition of Caries
  ∘ Caries is defined by the occurrence of a restoration, endodontic procedure, oral surgery or a prosthodontic procedure.
  ∘ CDT codes: D2000- D3999, D5000 - D7999

### MEPS (Survey)

- Enrolled Population
  ∘ Included all respondents answered the following question in affirmative:
    ▪ Ever had Medicaid/SCHIP during 2018?
- Dental Patients
  ∘ Respondents are dental patients if their response to the following question was one or more:
    ▪ Number of dental care visits 2018
- Caries Treatment
  ∘ Respondents are considered to have a caries treatment if they answered any of the following questions in the affirmative:
    ▪ Restorations: Fillings, Inlays, Crowns or Caps
    ▪ Root canals
    ▪ Prosthodontics: Bridges, dentures, reliners/repair bridges/denture repairs or implants
    ▪ Extractions: tooth pulled or other oral surgery
- Periodontics: defined as individuals having any periodontal scaling, root planning or gum surgery in 2018

## Appendix 2: Unweighted Survey Data (MEPS)

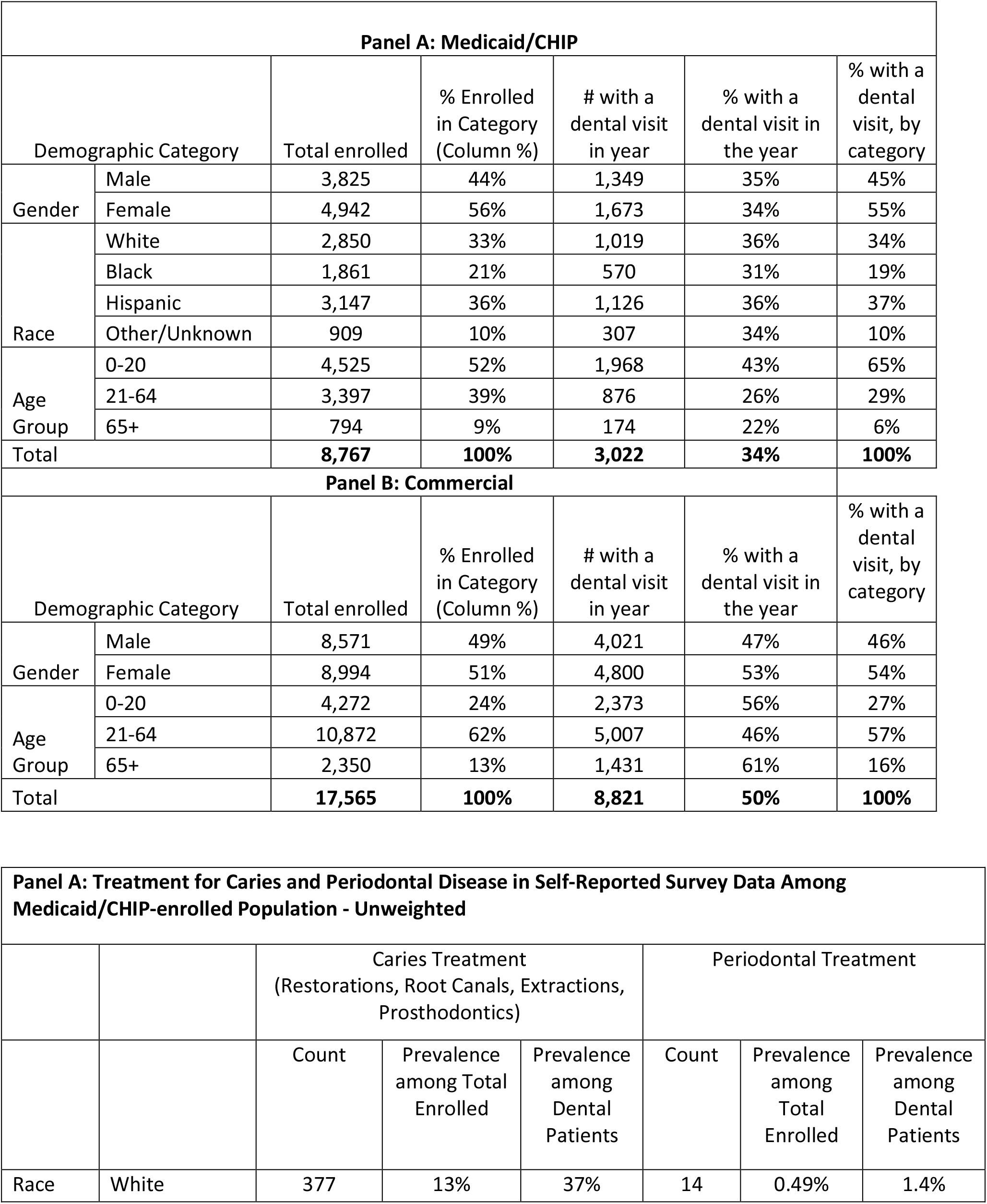
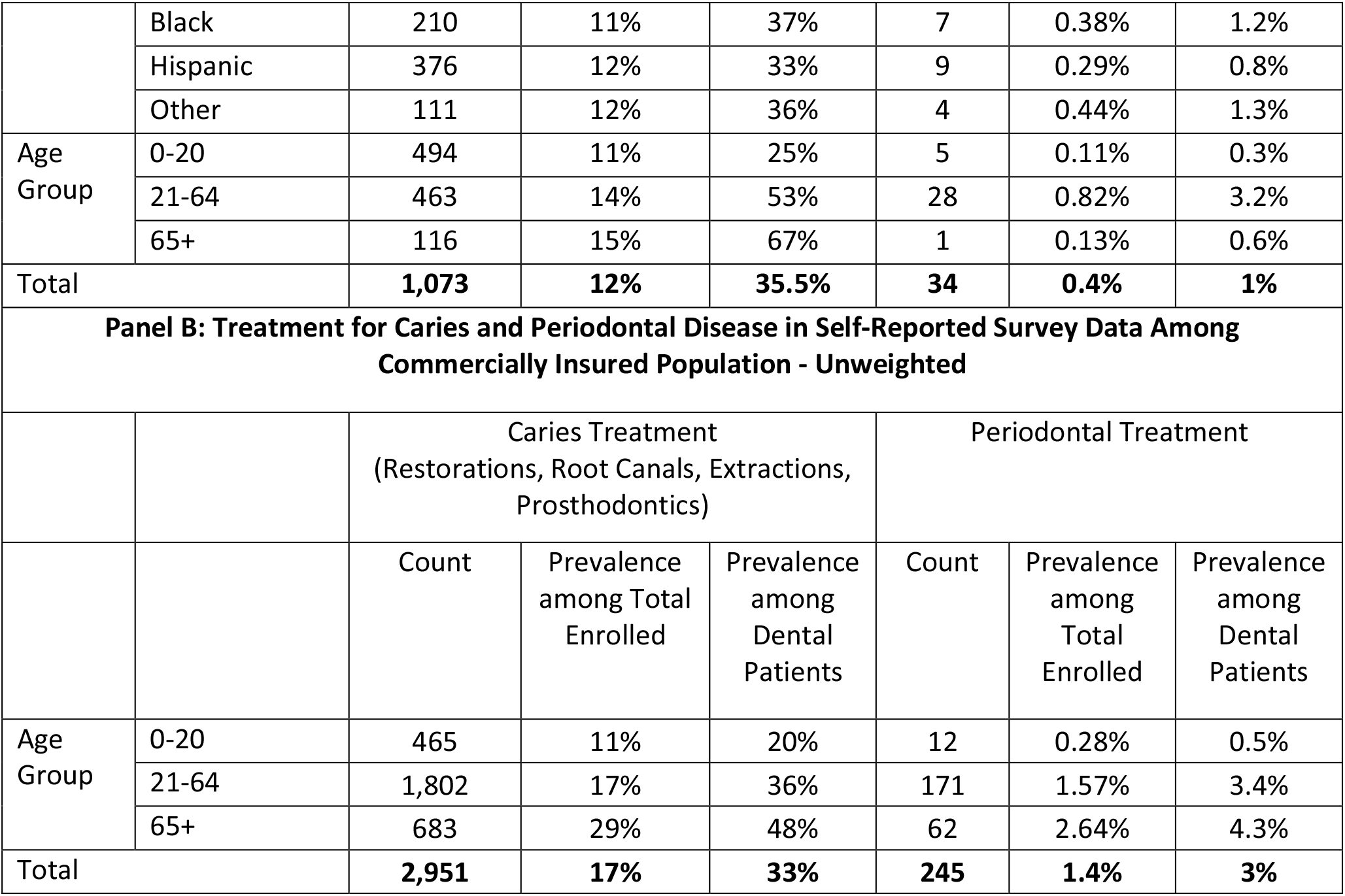

## Notes

### Competing Interest Statement

The authors have declared no competing interest.

### Funding Statement

This study received no external funding.

### Author Declarations

This study received IRB exemption from the Western Institutional Review Board.

### Summary of Updates

Updated the authors.

## Reference

1. Wilson J. Bock A. The benefit of using both claims’ data and electronic medical record data in health care analysis. OptumInsight White paper. Available at: file:///E:/DentalQuest/Resources/Benefits-of-using-both-claims-and-EMR-data-in-HC-analysis-WhitePaper-ACS.pdf. Accessed April 30, 2020

2. Broder MS, Cai B, Chang E, Neary MP. Incidence and prevalence of neuroendocrine tumors of the lung: analysis of a US commercial insurance claims database. BMC Pulmonary medicine 2018: 18: 135.

3. Furst DE, Clarke AE, Fernandes AW, Bancroft T, Greth W, Iorga SR. Incidence and prevalence of adult systemic lupus erythematosus in a large U.U. managed-care population. Lupus 2013; 22: 99–105

4. Cragin LA, Laney AS, Lohff CJ, Martin B, Pandiani JA, Blevins LZ. Use of insurance claims data to determine prevalence and confirm a cluster of sarcoidosis cases in Vermont. Public Health Report 2009; 124: 442–446.

5. Goldner EM, Jones W, Waraich P. Using administrative data to analyze the prevalence and distribution of schizophrenic disoeders. Psychiatric services, 2003:54(7); 1017–1021.

6. Cooper GS1, Yuan Z, Jethva RN, Rimm AA. Use of Medicare claims data to measure county-level variation in breast carcinoma incidence and mammography rates. Cancer Detect Prev. 2002;26(3):197–202.

7. Sloan FA, Derek SB, Carlisle ES, Ostermann J, Lee PP. Estimation of incidence rates with longitudinal claims data. Arch Ophthamol 2003; 121:1462–1468.

8. Tessier-Sherman, B., Galusha, D., Taiwo, O.A. et al. Further validation that claims data are a useful tool for epidemiologic research on hypertension. BMC Public Health 13, 51 (2013). https://doi.org/10.1186/1471-2458-13-51

9. Okunseri CE, Okunseri E, Garcia RI, Visotcky A, Szabo A Geographic variations in dental sealant utilization by Medicaid enrollees..J Public Health Dent. 2020 Nov 11.

10. Okunseri C, Zbin S, Zheng C, Eichmiller F, Okunseri E, Szabo A.Emergency department visits and dental procedures: Mission of Mercy, 2013-2016. Clin Cosmet Investig Dent. 2019 Jun 13;11:157–162. doi: 10.2147/CCIDE.S207266. eCollection 2019.

11. Wagner K, Szabo A, Zheng C, Okunseri E, Okunseri C Billed and Paid Amounts for Preventive Procedures in Dental Medicaid..JDR Clin Trans Res. 2019 Oct;4(4):371–377

12. Li KY, Okunseri CE, McGrath C, Wong MC.Self-Reported General and Oral Health in Adults in the United States: NHANES 1999-2014. Clin Cosmet Investig Dent. 2019 Dec 24;11 :399–408

13. Chen X, Naorungroj S, Douglas CE, Beck JD. Self-reported Oral Health and Oral Health Behaviors in Older Adults in the Last Year of Life. J. Gerontology 2013; 68 (10): 1310–1315

14. National Association of Dental Plan Inc. 2012 NADP/DDPA joint Benefits Report :Enrollment. Available at: C Copy of Dental Insured Uninsured.xlsx (nadp.org) accessed on March 12, 2021.

15. . Fleming E, Afful J. Prevalence of total and untreated dental caries among youth: United states, 2015-2016. NCHS Data Brief, no 307. Hyattsville, MD: National Center for Health Statistics. 2018.

16. Broadbent JM, Thomson WM, Poulton R. Trajectory patterns of dental caries experience in the permanent dentition to the fourth decade of life. J Dent Res 2008; 87: 69–72.

17. Hyman J. The limitations of using insurance data for research. JADA 2015, 146 (5): 283–285.

